# Persistent SARS-CoV-2 infection and increasing viral variants in children and young adults with impaired humoral immunity

**DOI:** 10.1101/2021.02.27.21252099

**Authors:** Thao T. Truong, Alex Ryutov, Utsav Pandey, Rebecca Yee, Lior Goldberg, Deepa Bhojwani, Paibel Aguayo-Hiraldo, Benjamin A. Pinsky, Andrew Pekosz, Lishuang Shen, Scott D. Boyd, Oliver F. Wirz, Katharina Röltgen, Moiz Bootwalla, Dennis T. Maglinte, Dejerianne Ostrow, David Ruble, Jennifer H. Han, Jaclyn A. Biegel, Maggie Li ScM, ChunHong Huang, Mayala K. Sahoo, Pia S. Pannaraj, Maurice O’Gorman, Alexander R. Judkins, Xiaowu Gai, Jennifer Dien Bard

## Abstract

**Background:** There is increasing concern that persistent infection of SARS-CoV-2 within immunocompromised hosts could serve as a reservoir for mutation accumulation and subsequent emergence of novel strains with the potential to evade immune responses.

**Methods:** We describe three patients with acute lymphoblastic leukemia who were persistently positive for SARS-CoV-2 by real-time polymerase chain reaction. Viral viability from longitudinally-collected specimens was assessed. Whole-genome sequencing and serological studies were performed to measure viral evolution and evidence of immune escape.

**Findings:** We found compelling evidence of ongoing replication and infectivity for up to 162 days from initial positive by subgenomic RNA, single-stranded RNA, and viral culture analysis. Our results reveal a broad spectrum of infectivity, host immune responses, and accumulation of mutations, some with the potential for immune escape.

**Interpretation:** Our results highlight the need to reassess infection control precautions in the management and care of immunocompromised patients. Routine surveillance of mutations and evaluation of their potential impact on viral transmission and immune escape should be considered.

**Funding:** The work was partially funded by The Saban Research Institute at Children’s Hospital Los Angeles intramural support for COVID-19 Directed Research (X.G. and J.D.B.), the Johns Hopkins Center of Excellence in Influenza Research and Surveillance HHSN272201400007C (A.P.), NIH/NIAID R01AI127877 (S.D.B.), NIH/NIAID R01AI130398 (S.D.B.), NIH 1U54CA260517 (S.D.B.), an endowment to S.D.B. from the Crown Family Foundation, an Early Postdoc.Mobility Fellowship Stipend to O.F.W. from the Swiss National Science Foundation (SNSF), and a Coulter COVID-19 Rapid Response Award to S.D.B. L.G. is a SHARE Research Fellow in Pediatric Hematology-Oncology.

## Introduction

Infection with severe acute respiratory syndrome coronavirus-2 (SARS-CoV-2), the cause of coronavirus disease 2019 (COVID-19) is most commonly detected using real-time polymerase chain reaction (RT-PCR). Viral load typically peaks with onset of symptoms and wanes to undetectable levels by week three, when patients generally begin to develop antibodies.^1^ While the Centers for Disease Control and Prevention (CDC) recommends quarantining for 14 days following exposure to COVID-19, the time course of infection may vary due to factors including age, immune status, and disease severity.^2^ In most patients, culture, contact tracing, and subgenomic RNA detection studies have not demonstrated infectivity beyond 10 days of symptom onset.^3,4^ In rare cases prolonged shedding of SARS-CoV-2 has been observed in immunocompromised adults, making study of this population critically important.^5–8^ However, the temporal dynamics of SARS-CoV-2 infectivity and the evolution of SARS-CoV-2 mutational profiles over prolonged periods of infection in immunocompromised patients, particularly in children, have not been described. Beyond the implications for individual patients, the emergence of B.1.1.7 SARS-CoV-2 and other lineages with potential for immune evasion has led to greater focus on the importance of genomic surveillance.^9,10^ Here, we describe prolonged SARS-CoV-2 RT-PCR positivity in two children and one young adult undergoing therapy for B-cell acute lymphoblastic leukemia (ALL). In addition to evidence of ongoing replication, two of these cases demonstrated significant intra-host SARS-CoV-2 mutational accumulation and host immune responses that may have contributed to their disease course.

## Methods

### Case histories

Patient 1 is a previously healthy female under 5 years of age with no significant past medical history prior to several admissions to the emergency department (ED) for worsening pancytopenia, decreased appetite, and abdominal pain. Her bone marrow biopsy revealed 58% blasts consistent with B-cell ALL and a chemotherapy regimen was initiated (Figure 1A, Table 1). Asymptomatic screening for SARS-CoV-2 by RT-PCR at time of discharge revealed a positive result (day 0). She was re-admitted to the hospital on day 3 for fatigue and vomiting as well as cough, malaise, and gastrointestinal symptoms and discharged on day 6. She consistently tested positive for SARS-CoV-2 during follow-up screens for her chemotherapy until she finally tested negative on day 91 without any notable respiratory symptoms (Figure 1A, Figure 2A).

**Figure 1.**
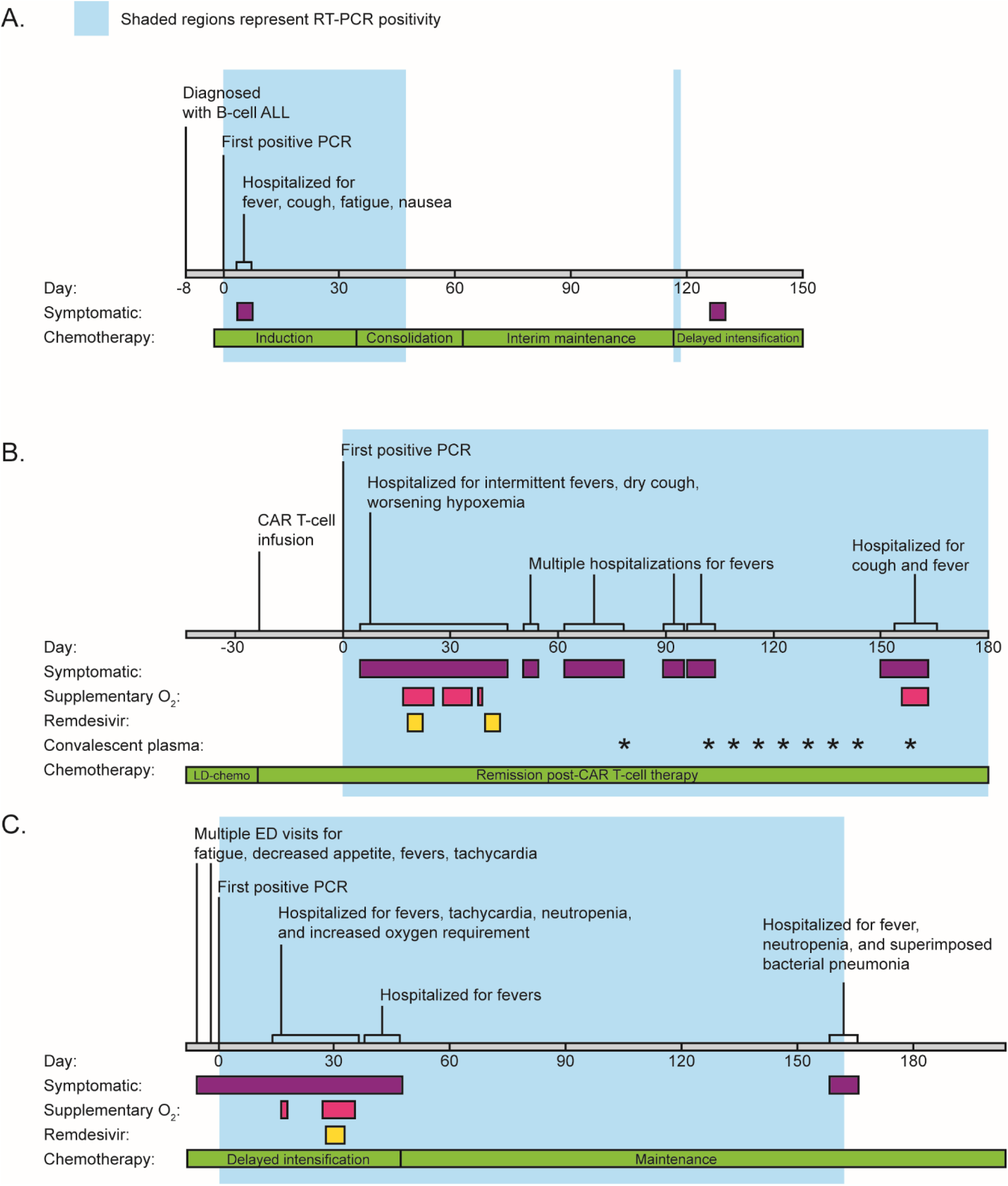
Clinical timeline of symptoms, hospital admissions, and treatment. Timelines for patient 1 (A), patient 2 (B), and patient 3 (C) are labelled by date from initial positive RT-PCR (day 0). Colored bars indicate time periods where patients were symptomatic, required supplementary oxygen, or received treatment (Remdesivir or convalescent plasma). The phases of chemotherapy are also shown.

**Table 1.** Demographic, oncological, and clinical characteristics of study patients.

|  | Patient 1 | Patient 2 | Patient 3 |
| --- | --- | --- | --- |
| **Demographic characteristics** |  |  |  |
| Age range (years) | 0-5 | 20-25 | 0-5 |
| Sex | Female | Male | Male |
| Body Mass Index | 16.6 kg/m<sup>2</sup> | 27 kg/m<sup>2</sup> | 16.3 kg/m<sup>2</sup> |
| **Oncological characteristics** |  |  |  |
| Oncological diagnosis | SR B-ALL | HR B-ALL, refractory | HR B-ALL |
| Other coexisting medical conditions | None | Obstructive sleep apnea<br>Hypothyroidism<br>Intermittent Asthma | None |
| Oncological treatment | Chemotherapy | Lymphodepleting chemotherapy and CAR-T cell therapy | Chemotherapy |
| Phases of oncological treatment during viral shedding | -Induction<br>-Consolidation<br>-Interim maintenance<br>-Delayed intensification | Post- CAR-T cell therapy | -Delayed intensification<br>-Maintenance |
| Active immunosuppressive or immunomodulatory drugs? | SR B-ALL chemotherapy per COG AALL0932: Dexamethasone, vincristine, peg-asparaginase, methotrexate, 6-MP, doxorubicin | Cyclophosphamide and Fludarabine as lymphodepleting agents prior to CAR-T cell therapy | HR B-ALL chemotherapy per COG AALL0232: Cyclophosphamide, cytarabine, thioguanine, vincristine, dexamethasone, methotrexate, mercaptopurine |
| Chemotherapy | Yes (detailed above) | Yes (detailed above) | Yes (detailed above) |
| Corticosteroids | Yes (per chemotherapy protocol) | No | Yes (per chemotherapy protocol) |
| CAR-T therapy | No | Yes, Tisagenlecleucel | No |
| **Clinical, laboratory and imaging data** |  |  |  |
| Days of PCR positivity | 46 (inconclusive/positive again day 116-117) | 165+ from time of report | 162 |
| Symptomatic at time of initial positivity | No | No | Yes |
| Symptomatic at any positive time point | Yes | Yes | Yes |
| **Symptoms** |  |  |  |
| Fever | Yes | Yes | Yes |
| Malaise | Yes | Yes | Yes |
| Cough | Yes | Yes | Yes |
| Sore throat | Yes | No | Yes |
| Headache | No | No | No |
| Shortness of breath | No | Yes | Yes |
| Congestion/rhinorrhea | No | Yes | Yes |
| Vomiting | Yes | Yes | Yes |
| Abdominal pain | Yes | Yes | Yes |
| Anosmia and Ageusia | Unable to assess | Unable to assess | Unable to assess |
| Other infectious disease findings | No | - *Clostridioides difficile* diarrhea<br>- *Staphylococcus hominis* bacteremia (day +7) | Superimposed bacterial pneumonia (day 162, improved with antibiotics) |
| Laboratory data at initial positivity | WBC: 2.14 K/μL<br>ANC: 0.74 K/μL<br>ALC: 1.36 K/μL | WBC: 2.30 K/μL<br>ANC: 1.59 K/μL<br>ALC: 0.42 K/μL | WBC: 0.65 K/μL<br>ANC: 0.00 K/μL<br>ALC: 0.56 K/μL |
| Lymphocyte subsets | Not done | Day +7 to +158 (multiple)<br>CD3: 75-1000 cells/μL<br>CD4: 31-314 cells/μL<br>CD8: 52-747 cells/μL<br>CD19: <1 cell/μL throughout | Day +13<br>CD3: 1204 cells/μL<br>CD4: 194 cells/μL<br>CD8: 987 cells/μL<br>CD19: 1 cells/μL |
| Chest CT scan | Not done | Bilateral patchy ground glass opacities involving all lobes, predominantly peripherally in lower lobes (similar in all 3 CT scans on days +38, +73 and +116) | Persistent multifocal pneumonia and scattered areas of ground-glass opacities, with shifting opacities increased in some areas but improved in others (CT scans on days +22, +47, +96, +166) |
| **Treatment for COVID-19 infection** |  |  |  |
| Oxygen supplementation | No | Yes, intermittent (maximal 10L per minute via facemask) | Yes (maximal 5L per minute via facemask) |
| COVID-19 related ICU admissions | No | Yes (5 days) | No |
| Mechanical ventilation | No | No | No |
| Remdesivir | No | Yes, two 5 day courses | Yes, one 5 day course |
| Convalescent plasma | No | Yes (measured as ratio serum to calibrator absorbance)<br>Day 78: 4.3<br>Day 103: 9.1<br>Day 110: 8.6<br>Day 123: 8.3<br>Day 130: 7.5<br>Day 137: 4.6<br>Day 144: 1.4<br>Day 158: 4.1<br>Day 165: 3.4<br>Day 172: 1.53 | No |

SR, standard risk; HR, high risk; B-ALL, B-cell acute lymphocytic leukemia; CAR-T, chimeric antigen receptor T-cell therapy; COG, Children's Oncology Group; WBC, white blood cells; ANC, absolute neutrophil count; ALC, absolute lymphocyte count; CT, computerized tomography; ICU, intensive care unit

**Figure 2.**
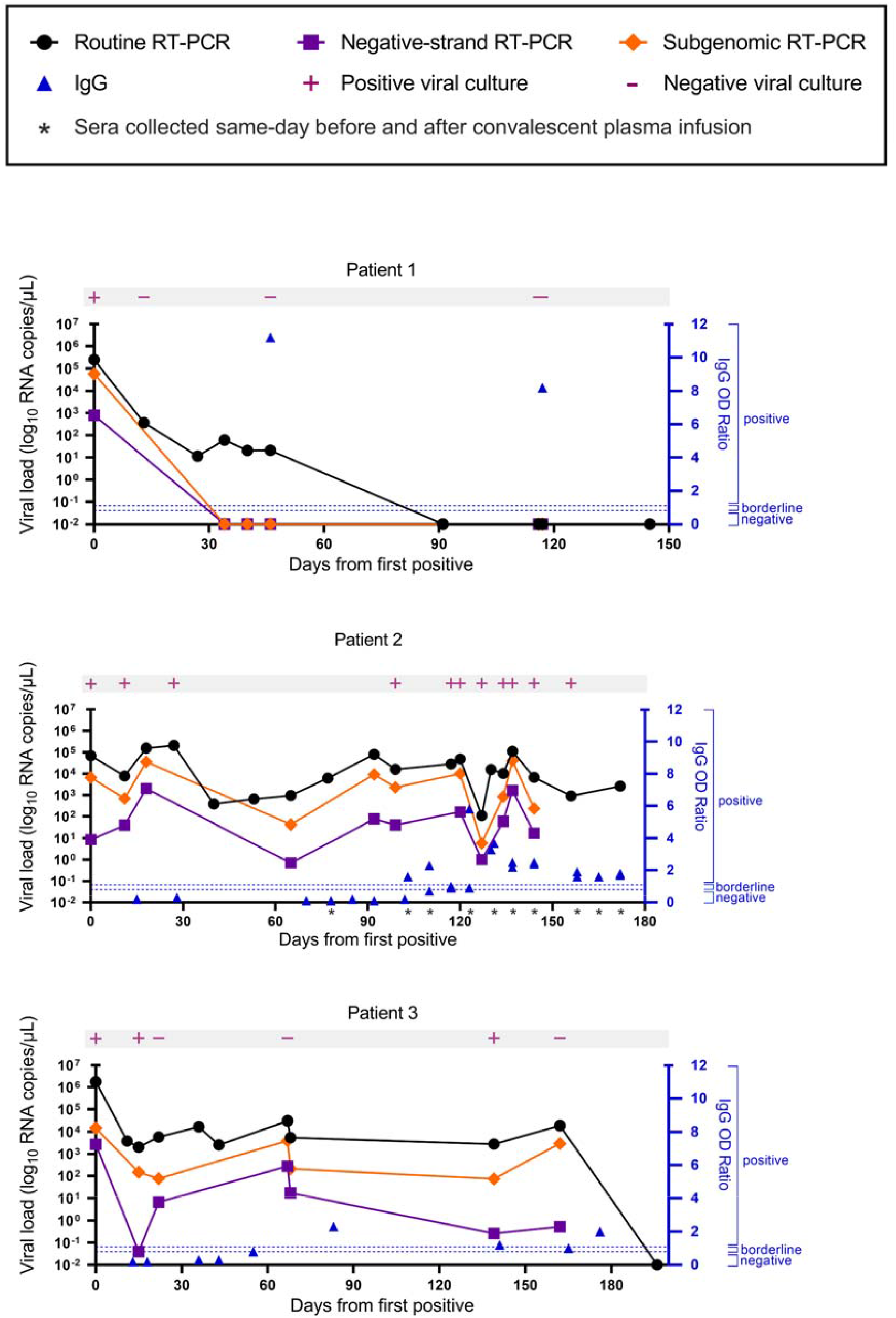
Viral load by routine, negative-strand, and subgenomic RT-PCR. Time course of viral load from nasopharyngeal or combined nares/oropharyngeal swabs collected from each patient. Viral culture results are indicated in pink. Corresponding serum anti-SARS-CoV-2 IgG values are plotted in blue. For patient 2, IgG was measured before and after administration of convalescent plasma at the indicated timepoints.

Patient 2 is a male in the 20-25 year age range who was previously diagnosed with B-cell ALL six months prior to his first SARS-CoV-2 positive RT-PCR. He received combination chemotherapy, however continued to have detectable residual disease at the end of the consolidation phase, prompting initation of CD19-directed chimeric antigen receptor T cell therapy (CAR-T, Tisagenlecleucel, Cassiopeia protocol). Upon a follow up visit to the transplant and cell therapy clinic 3 weeks later, the patient was tested for SARS-CoV-2 due to concerns for possible contact exposure and was positive (day 0). On day 6 after the initial positive test he presented to the ED with decreased appetite, fever, and two days of dry cough. His chest x-ray showed a diffuse interstitial prominence and a small bilateral pleural effusion. He developed intermittent fevers with increased oxygen requirement over the next several days, including an episode of acute hypoxemic respiratory failure necessitating 10 liters of oxygen via face mask on day 18. He received a 5-day course of Remdesivir. From then on his oxygen requirement was weaned down to 2-3 liters via nasal cannula (NC), however he continued to have intermittent fevers and bilateral patchy ground glass opacities on chest CT. He improved after a second 5-day course of Remdesivir, weaned down off supplemental O2 to his nighttime baseline of 0.5L/NC while asleep (used to treat his mild obstructive sleep apnea) and was discharged on day 45. He was hospitalized four additional times for persistent fevers with negative infectious work-up except for a persistently positive SARS-CoV-2 RT-PCR with consistently high (Ct range: 17.2 – 28.1) viral loads. Weekly convalescent plasma therapy was initiated on day 103 and changed to biweekly beginning day 144. Unfortunately on day 156, he developed worsening cough, new fevers, and was found to be hypoxemic down to 80% needing supplemental oxygen with 4L/NC. Full infectious disease workup failed to demonstrate any additional infection besides persistently positive SARS-CoV-2 RT-PCR. He was admitted to the hospital and received convalescent plasma. Thereafter, he resumed weekly convalescent plasma infusions. His anti-SARS-CoV-2 IgG was finally positive on day 103 which was likely due to the plasma infusion. He has remained SARS-CoV-2 RT-PCR positive through his most recent test on day 250 at the time of this publication.

Patient 3 is a male under 5 years of age who was diagnosed with high-risk B-cell ALL 7 months prior to presentation to the ED with fever and confirmed positive for SARS-CoV-2 upon admission (day 0). At admission he demonstated tachycardia and pancytopenia that were attributed to his chemotherapy. After clinical improvement, he was discharged on day 6 but re-admitted on day 14 with recurrent fever and tachycardia. He continued to be febrile, had worsening pancytopenia, pulmonary infiltrates, and increasing oxygen requirements to 5L via face mask by day 32, at which time he received a 5-day regimen of Remdesivir. His fevers and respiratory distress improved and he was discharged on day 40, only to be readmitted for fever on day 43 and discharged on day 51. The patient then started maintenance chemotherapy. Anti-SARS-CoV-2 IgG was repeatedly negative or borderline and finally became positive on day 83. He was re-admitted on day 162 for possible urinary tract infection and subsequently developed tachypnea, fever, and cytopenia. Chest CT scans on days 47, 96, and 166 showed persistent multifocal pneumonia and scattered areas of ground-glass and consolidative opacities, with shifting opacities that increased in some areas but improved in others. He improved on antibiotics and was discharged on resolution of his fevers on day 173. He was last RT-PCR positive on day 162 and finally negative on day 196. A surveillance chest CT scan on day 230 showed near complete resolution of his pulmonary disease.

### Study cohort

Specimens from all three patients were obtained at Children’s Hospital Los Angeles (CHLA) between May 7 to November 21, 2020 for clinical testing for various indications including hospital admissions both related and unrelated to COVID-19, asymptomatic pre-procedural screening, evaluation of COVID-19 related and unrelated symptoms, and after a high-risk exposure to someone with confirmed SARS-CoV-2 infection. Data for demographics, clinical course and management were obtained from the electronic medical record (EMR). Disease severity was categorized using Centers for Disease Control and Prevention’s classification as previously described.^11^ The study was approved by the CHLA Institutional Review Board (CHLA-16-00429).

### Detection of SARS-CoV-2 RNA

Nasopharyngeal swabs or combined oropharyngeal and nares swabs were sent to the Clinical Virology Laboratory at CHLA for testing. The molecular assays used were the CDC 2019-Novel Coronavirus Real-Time RT-PCR assay, the Taqpath COVID-19 RT-PCR assay (Thermo Fisher, Walham, MA), the Xpert Xpress SARS-CoV-2 assay (Cepheid, Sunnyvale, CA), and the Simplexa COVID-19 assay (Diasorin Molecular, Cypress, CA). Total nucleic acid was extracted according to manufacturer’s instructions as outlined in the respective Emergency Use Authorization.^11–14^ Viral loads (copies/mL) were calculated based on a standard curve generated by testing samples with known viral copy numbers for each assay as previously described.^11^

### Viral culture

Viral culture was performed using SARS-CoV-2-susceptible VeroE6TMPRSS2 cells^15,16^ adapted from the VeroE6 cell line (ATCC CRL-1586) to express the TMPRSS2 protease at levels approximately 10-fold higher than that found in the human lung. 150 µL of clinical specimen was used to inoculate one well of a 24 well plate of VeroE6TMPRSS2 cells as previously described.^15,16^ After a 2-hour incubation at 37°C, the inoculum was removed and replaced with 0.5 ml infection media and the plates incubated at 37°C for 5 days. The plates were scored daily for cytopathic effect and supernatants harvested when >75% of the cells had detached. The presence of SARS-CoV-2 viruses in the harvested supernatants was then verified by quantitative RT-PCR using the CDC N1 and RNase P primer sets on RNA extracted with the viral RNA isolation kit (Qiagen, Germantown, MD).

### Quantitative Two-step strand-specific SARS-CoV-2 RT-PCR

A two-step strand-specific SARS-CoV-2 RT-PCR assay targeting the *envelope* (E) gene was performed as previously described by Hogan et al. as a biomarker to predict presence of actively replicating virus.^17^ In the first set of reactions, reverse transcription with strand-specific primers converts SARS-CoV-2 RNA to complementary DNA (cDNA). In the second step, the cDNA is amplified by real-time PCR in using the Rotor-Gene Q instrument (QIAGEN). A standard-curve to convert minus-strand Ct values to copies/µL was generated using *in vitro* transcribed minus-strand *E* gene RNA.

### Subgenomic SARS-CoV-2 RT-PCR

One-step subgenomic (sg) SARS-CoV-2 RT-PCR utilizing a forward primer targeting the 5’ leader sequence and reverse primer and probe complementary to *E* gene sequences was adapted from Wolfel et al. and combined in multiplex with the Hong Kong Orf1ab and Centers for Disease Control and Prevention RNAse P primer/probe sets^18,19^ as an additional biomarker for actively replicating virus. Real-time RT-PCR was performed using the Rotor-Gene Q instrument. A standard-curve to convert sgRNA Ct values to copies/µL was generated using *in vitro* transcribed plus-strand RNA encoding the leader and *E* gene sequences (Log_10_ copies/µL = −0.288*Ct + 11.351).

### Measurement of anti-SARS-CoV-2 antibodies in serum

IgG to the S1 domain of SARS-CoV-2 spike was measured from serum within 24 hours of collection with a commercial enzyme-linked immunosorbent assay (ELISA) kit (EUROIMMUN, Lubeck, Germany).^20^ A ratio of sample to calibrator optical density of < 0.8 is considered negative, ≥ 0.8 to <1.1 is considered borderline, and ≥1.1 is considered positive.

In addition, IgA, IgG and IgM to SARS-CoV-2 nucleocapsid (N) protein, spike S1, and receptor binding domain (RBD) and the competition ELISA procedure were measured as recently described by Röltgen et al.^21^

### Viral genome library construction and sequencing

Whole-genome sequencing (WGS) libraries of viral cDNA were prepared as previously described using the CleanPlex SARS-CoV-2 Research and Surveillance NGS Panel (Paragon Genomics, Hayward, CA).^11^ Libraries were then quantified and normalized to approximately 7nM and pooled to a final concentration of 4nM; libraries were denatured and diluted according to Illumina protocols and loaded on the MiSeq at 10pM. Paired-end and dual-indexed 2×150bp sequencing was done using Micro Kit v2 (300 Cycles). Libraries yielding average depth ≥ 1000x were considered high quality and used for downstream analysis, and variant calling was performed for nucleotide positions with ≥100x coverage.

### Genome assembly and variant calling

Coverage profiles, variant calls and consensus genomes were generated using the in-house software system LUBA (Lightweight Utilities for Bioinformatics Analysis).^11^ Ion Torrent aligner TMAP (Thermo Fisher) was used to generate AmpliSeq BAMs. Sequencing of majority of clinical specimens by two alternate methods were also pursued to validate Paragon variant calls (appendix p 1-3). Paragon and Twist nucleotide sequences were aligned with NovoAlign (Novocraft Technologies, Selengor, Malaysia) aligner. Ion Torrent aligner TMAP (Thermo Fisher) was used to generate AmpliSeq BAMs.

## Results

### Viral detection by RT-PCR and culture

Specimens from all three patients were collected and used to detect SARS-CoV-2 RNA over the course of 6 months. In addition to viral culture, we performed strand-specific and subgenomic RT-PCR to detect ongoing replication.^4,17,18^ The viral load in patient 1 was highest at day 0 and then gradually declined and was last reliably detected on day 46 (Figure 2A). Culturable virus was only detected in the day 0 sample and subsequent cultures were negative. The SARS-CoV-2 minus strand and subgenomic RNA was only detected from the first sample taken on day 0 and not in any subsequent samples (Figure 2A).

The viral load in patient 2 remained consistent through day 172 despite two courses of Remdesivir and weekly administration of convalescent plasma. Consistent with these results, SARS-CoV-2 was recovered by viral culture for up to day 144. Viral cultures were not explored past 144 days from initial positive. Strikingly, the minus strand and subgenomic RNA was detected in nearly all of the samples from patient 2 and the Ct values trended with the corresponding viral load (Figure 2B).

Patient 3 also maintained consistent viral loads through day 162 despite weak seroconversion on day 83, and finally tested negative on day 196 (Figure 2C). He demonstrated variable viral recovery with positive cultures from days 0, 22, and 139, and negative cultures from days 15, 67, and 162 which we believe reflects poor sample integrity or a transient reduction in infectious virus production given that subgenomic and minus strand RNA was readily detectable up through day 162.

### Antibody response

We assessed each patient’s serological response profile using ELISAs to measure IgA, IgG, and IgM antibodies specific to the SARS-CoV-2 receptor binding domain (RBD), S1, and N proteins. Patient 1 had a strong IgA, IgG, and IgM response to all tested antigens at days 46 and 117, consistent with their more typical clinical course of infection and recovery, with viral RNA not reliably detected after day 46 (Figure 3A). In contrast, patient 2, who had culturable virus up to day 144 and positive viral RNA up to day 172, was more weakly IgG positive and was negative for IgA and IgM to all tested SARS-CoV-2 antigens from days 110-144 (Figure 3B). Importantly, paired sera obtained from patient 2 before and after convalescent plasma infusion showed increased IgG levels after infusion which were rapidly waning. As this patient had B-cell aplasia following CAR T-cell therapy, his ability to mount an immune response was impaired. Patient 3 had detectable viral RNA at least up to day 162, and showed serological results spanning days 83-176 with a weakly positive IgG and unusually high levels of IgM for the RBD, S1, and Spike proteins, but negative N antibodies (Figure 3C). Antibody levels were compared to four non-immunocompromised COVID-19 patients, who showed the typical increase of anti-SARS-CoV-2 IgG, IgM, and IgA antibodies between one to two weeks after symptom onset. IgA and IgM antibody levels began to wane about 3 to 4 weeks after symptom onset, while IgG levels were more stable (Figure 3D).

**Figure 3.**
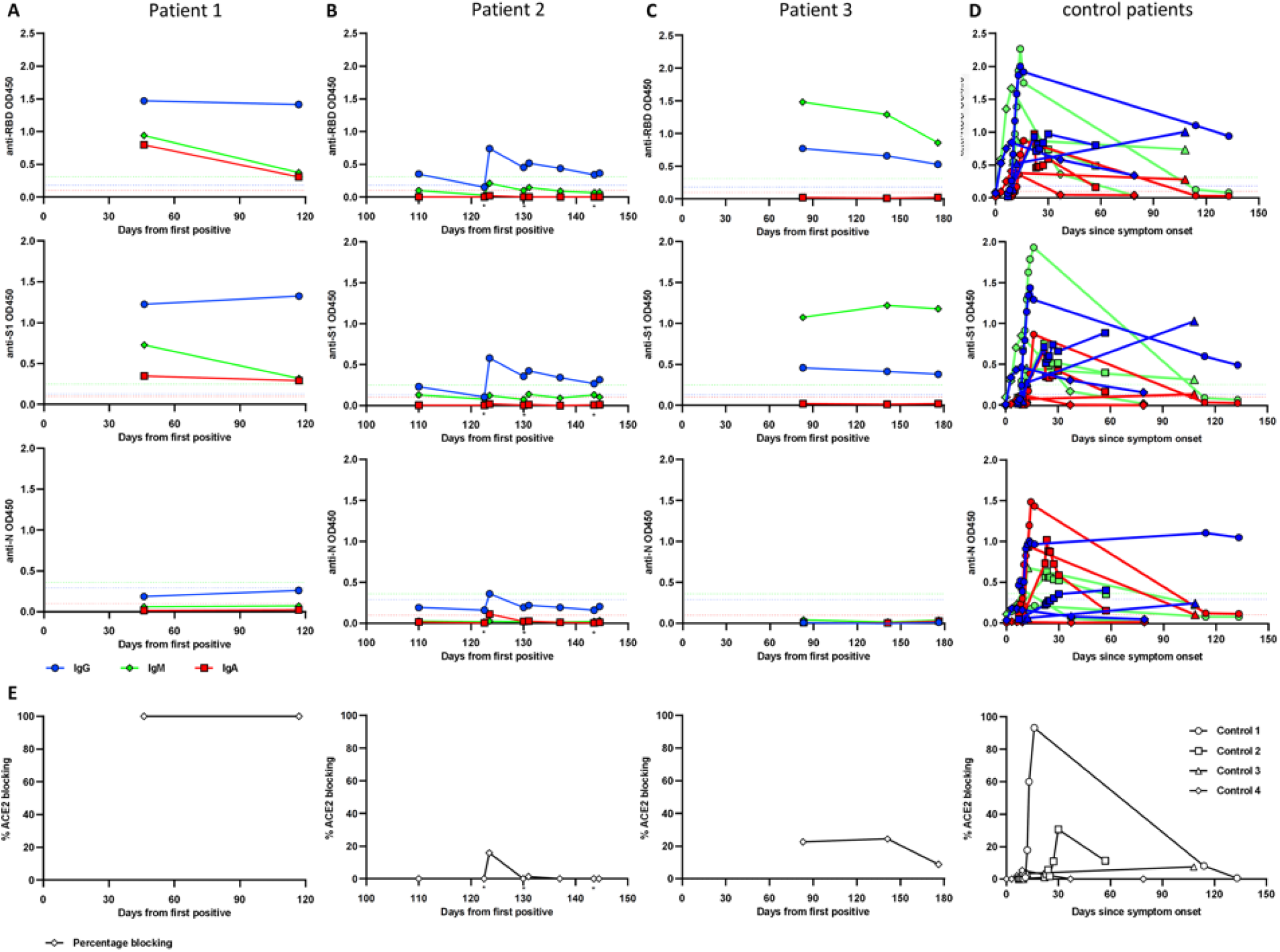
Anti-SARS-CoV-2 antibody responses in three immunocompromised pediatric and young adult patients. Serum samples from two immunocompromised children (A, C) and one young adult (B) were collected up to 176 days after initial positive SARS-CoV-2 PCR test. Plasma samples from four non-immunocompromised COVID-19 patients were analyzed for comparison (D). Antibodies specific for SARS-CoV-2 Spike RBD (top row), S1 subunit (second row), and N protein (third row) were measured using ELISA at 1:100 serum/plasma dilution. RBD-ACE2 blocking activity was assessed in a competition ELISA and is shwon as percentage of blocking (E). The cutoff for seropositivity was defined as the mean absorbance + 3 SD of 94 pre-pandemic plasma samples in each assay (A, B, C, D). Color coding for isotypes: IgG (blue), IgM (green), IgA (red). Dotted lines depict the cutoff for seroconversion for the different isotypes in each assay. Individual donors were shown with different symbols (D). * indicates timepoints of convalescent plasma infusion in patient 2.

We also measured the ability of the patients’ sera to interfere with the interaction between the RBD of the SARS-CoV-2 spike protein and the angiotensin-converting enzyme 2 (ACE2) receptor *in vitro* (Figure 3E). Sera from patient 1 from days 46 and 117 achieved 100% blocking, whereas patient 2’s sera only weakly blocked the ACE2-RBD interaction on day 123 post-infusion and the other timepoints failed to block the interaction. Samples from patient 3 from days 83, 141, and 176 also showed poor blocking activity (Figure 3E). Plasma samples from the four control patients showed varying degrees of RBD-ACE2 blocking activity. In a previous study higher RBD-ACE2 blocking percentages were correlated with higher anti-RBD antibody levels^21^ The antibody results were overall consistent with what we observed from the blocking assay, with the strongest immune response in patient 1, least effective response in patient 2, and a weak response in patient 3.

### Genomic sequencing analysis

Specimens from patient 1 showed only a single major allele variant difference between days 0 and 27. Decreasing viral load precluded sequencing of specimens from later timepoints. Patients 2 and 3 demonstrated an increase in intra-host viral diversity over time in variants of both high (major) and low (minor) allele frequencies across the viral genome (Figure 4). In patient 2, 3 major allele variants emerged between days 0 and 40 with an additional 4 major and 7 minor allele variants by day 144. In patient 3, 13 major and 8 minor allele variants emerged by day 162. Virus clade predictions did not change across the specimens from each patient (patients 1 and 3: clade 20C and patient 2: clade 20A), providing further support that these patients were continuously infected rather than reinfected. Of particular note was the emergence of several spike gene mutations including 3 inframe deletions at residues S:Δ141-143, S:Δ145, and S:Δ141-144, a deletion-insertion at S:Δ211-212, and several nonsynonymous mutations including N440K, V483A, and E484Q at residues that have independently mutated in other lineages (Figure 4, appendix p 6-12). Sequencing of additional library preparations by two alternate methods confirmed the variant calls (appendix p 1-3).

**Figure 4.**
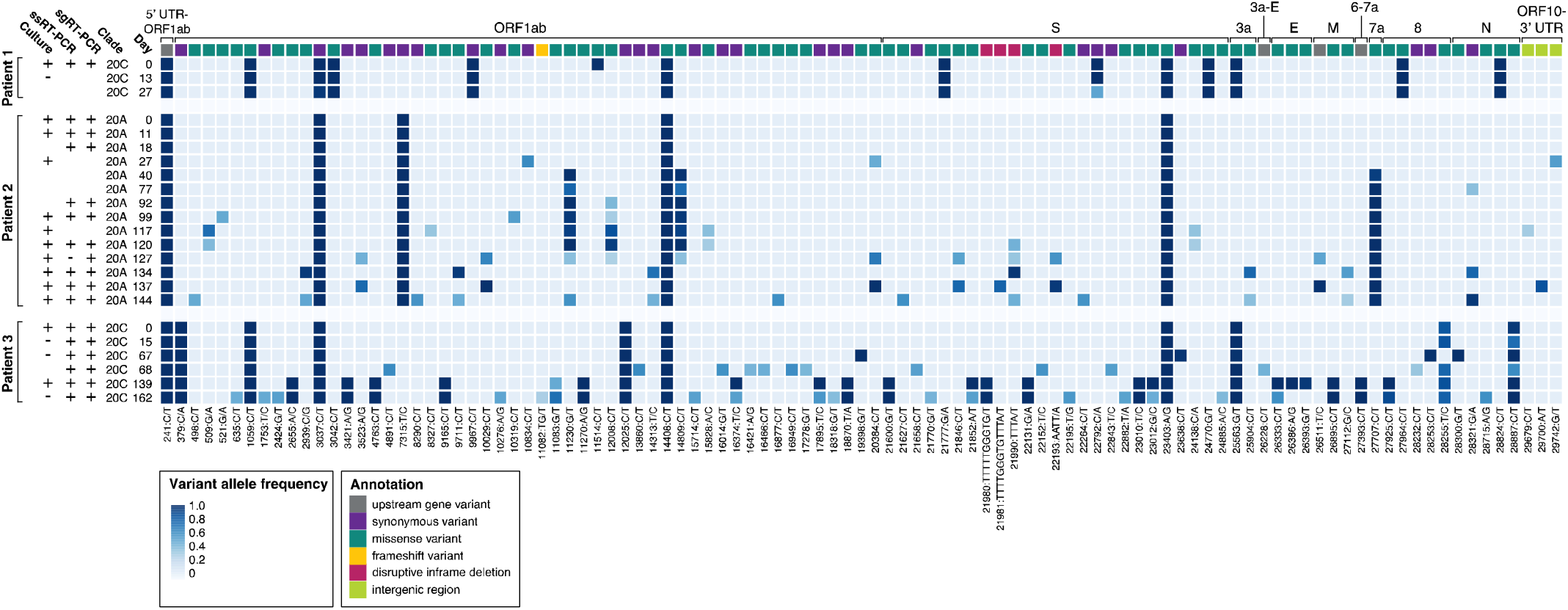
Accumulation of SARS-CoV-2 variants in three persistently positive pediatric and young adult patients with hematologic malignancies. Each row represents viral culture, strand-specific RT-PCR (ssRT-PCR), subgenomic RT-PCR (sgRT-PCR) and sequencing from longitudinally derived specimens numbered by days from initial positive test (day 0). Boxes represent distinct variants, and shading reflects variant allele frequency (cutoff = 0.25). Corresponding genes are labeled in top row and colored to represent variant annotation (see legend). Specimens from two patients demonstrate progressive accumulation of multiple variants and variants of lower allele frequencies (Patient 2, difference of 7 major allele variants between days 0 and 144; Patient 3, difference of 13 major allele variants between days 0 and 162). In addition to point mutations, four different inframe deletions were observed to develop in the S gene in Patient 2 and Patient 3. Several variants (both major and minor alleles) reverted at later timepoints. UTR, untranslated region; S, spike; E, envelope; M, matrix; N, nucleocapsid

## Discussion

We report two pediatric and one young adult patient with B-cell ALL with remarkably different courses of SARS-CoV-2 infection. Patient 1 had milder symptoms with rapidly declining and nonviable virus and serological evidence of a robust immune response that is representative of a typical COVID-19 clinical course.^1^ By contrast, patients 2 and 3 experienced more severe COVID-19 characterized by serological evidence of a weaker immune response with evidence of prolonged infectious virus shedding and mutation accumulation demonstrating emergence of potential escape mutations.

The infection control implications make it critically important to closely monitor immunocompromised patients who are persistently positive by SARS-CoV-2 RT-PCR and to determine whether or not the virus is actively replicating for a prolonged period of time. Using RT-PCR to detect subgenomic and negative-strand replication intermediates, we were able to identify ongoing replication throughout the samples from patient 2 (up to 144 days) and patient 3 (up to 162 days). It is notable that subgenomic RNA was readily detected for this length of time given that it is rarely detected past 8 days.^4,5,18^ Overall the detection of viral intermediates correlated very well with the viral culture data, suggesting these may serve an expeditious molecular surrogate for infectivity. Viral culture was intermittently positive up to day 139 in patient 3. As culturing methods are limited by sample quality and lower sensitivity compared to molecular methods, it is more likely that the consistently positive results by subgenomic and negative-strand RT-PCR accurately represented continued replication within patient 3.

There has been considerable interest in the rapidly spreading SARS-CoV-2 B.1.1.7 variant due to its potential increased transmissibility. Relative to the SARS-CoV-2 reference sequence, B.1.1.7 has acquired 17 mutations, 8 of which are in the spike gene. There is concern that such a divergent variant could have emerged from long-term replication within an immunocompromised host, especially with the lack of closely related viral isolates.^9^ Although we did not find these mutations in our patients, we did observe mutations in several regions within the spike gene. A V70P mutation appeared at day 162 in patient 3, overlapping the region where a S:Δ69-70del in the B.1.1.7 variant may enhance infectivity.^22^ Several other observed mutations mapped to important residues in the spike gene. We observed emergence of a S:Δ141-144 and S:Δ145 deletion in patient 2 and a S:Δ141-143 deletion in patient 3. Similar mutations in this region of the spike gene have been found to abolish the binding of the anti-spike protein 4a8 blocking/neutralizing monoclonal antibody (Figure 3, Tables S3-4).^7,23^ Additionally, in patient 3, N440K within the spike RBD emerged with an allele frequency of 0.51 on day 162 (Figure 3, Table S4). This variant was also found by another group to confer antibody escape.^24^ This mutation is adjacent to the position of a N439K variant of interest currently circulating in several lineages in Europe, which may enhance affinity of the binding of the spike protein to the ACE2 receptor and influences binding of monoclonal antibodies.^25^ At day 139 the sequenced specimen from patient 3 also had mutations at V483A and E484Q within the RBD. These residues were similarly associated with *in vitro* antibody binding and escape, and there has been interest in another mutation at the same position, E484K, which has emerged in the S.501Y.V2 lineage in South Africa (Figure 3, Table S4).^24,26–28^ Similar mutations at these positions (N440D, E484A, and E484K) have independently arisen within other persistently infected, immunocompromised patients.^5,7^

Analysis of patient antibody responses was performed to aid in understanding the persistent detection of SARS-CoV-2. Patient 1 was associated with very limited detection of active replication and had high levels of ACE2-RBD blocking antibodies, a surrogate measure of potential viral neutralizing antibodies *in vivo*. Patients 2 and 3 by contrast both had poor antibody responses, associated with evidence of prolonged infectivity. In immunocompetent patients, IgM, IgG, and IgA antibodies are generally detectable by day 20 post symptom onset.^21^ Patient 3 had an atypical persistence of high IgM levels on days 83, 141, and 176 without evidence of evolution to IgG or IgA, suggestive of an impaired immune response perhaps related to his partially immunocompromised state. It is unlikely that the detection of IgM in this patient is related to infection with a new viral strain as the majority of the mutations that emerged in patient 3 are rarely found in the general population (Table S4). In general, all three patients had higher levels of antibodies targeting RBD and S1 compared to N, and this pattern has been previously observed in patients with milder illness compared to patients who died or were admitted to intensive care units.^21^ However, compared to patients 1 and 2, patient 3 completely lacked antibodies to the N protein (Figure 3C). It is possible that the spike-focused immune response, which weakly blocked ACE2, led to the emergence of escape mutants. There were more mutations in the spike gene, particularly in the RBD region, in patient 3 compared to patient 2 and many of these emerged on day 139 or 162, a time period where his antibody levels were rising. Patient 2 was treated with two courses of remdesivir and several infusions of convalescent plasma, however antibody levels waned rapidly after each infusion, possibly due to redistribution in the body or partial consumption and clearance. These treatments failed to control viral replication and did not apparently lead to as many mutations with the potential for immune escape as observed in patient 3. We note that anti-SARS-CoV-2 antibody levels start to decrease as soon as one month after symptom onset in some previously healthy COVID-19 patients, therefore it is possible that our measurements at later time points may underestimate the peak antibody levels acheived by patients 2 and 3 throughout their clinical course.^21^

Our work demonstrates that immunocompromised pediatric and young adult patients are susceptible to prolonged viral infections with prolonged infectious virus shedding and mutation accumulation. The patients we report are immunocompromised to varying degrees, and support a potential correlation between host immune response and the emergence of viral variants, some of which may have the potential to escape antibody neutralization. One of the limitations of a small case series is the generalizability of findings and more work is needed to better understand the many host factors that can influence viral clearance and mutation within these patients. However, we conclude that the observed expanded intra-host heterogeneity of viral isolates from pediatric and young adult patients with suppressed immunity and correspondingly prolonged infection raises the possibility of emergence of variants with the potential to evade immune responses elicited during the course of infection or induced by vaccine. This possibility warrants further study and, in the meantime, serious consideration of extended infection control precautions and genomic monitoring in this patient population.

## Data Availability

The manuscript contains all relevant data.

## Acknowledgments

The authors thank the ATUM Bio team for optimization, production, and sharing of antigens used in this study. We would like to acknowledge NCBI, GISAID, and Nextstrain for providing valuable resources for SARS-CoV-2 genomics. We thank the National Institute of Infectious Diseases, Japan, for providing VeroE6TMPRSS2 cells.

## Research in context

### Evidence before this study

There has been substantial interest in the phenomenon of prolonged shedding of SARS-CoV-2. Although an increasing number of case reports have described the course of disease in immunocompromised individuals, definitive evidence for prolonged active replication with infectious virus and corresponding immunological data has been scarce. We searched PubMed, bioRxiv, and medrxiv for these reports using the terms “immunocompromised,” “immunosuppressed,” “SARS-CoV-2,” “replication,” “shedding,” and “infectivity” for relevant articles published up until December 26, 2020. We found four articles that have utilized longitudinal sequencing, detection of subgenomic RNA, and/or viral culture to demonstrate ongoing infectivity in immunocompromised adults.

### Added value of this study

Our study presents a more complete picture of the spectrum of prolonged infection for up to 162 days, which is the longest reported to-date, in three immunocompromised patients using viral culture, detection of both subgenomic and single-stranded RNA, whole-genome sequencing, and characterization of host serological responses. Secondly, this is the first report, to our knowledge, characterizing prolonged infection in pediatric and young adult patients. Thirdly, we demonstrated the clear presence of intra-host viral heterogeneity as evidence of ongoing viral evolution within the hosts. We note two of our patients acquired several mutations in the gene encoding the spike protein at specific residues that may be important for antibody binding. Finally, we present a potential correlation between the host immune response and the emergence of mutations in the spike gene.

### Implications of all the available evidence

Given recent interest in the emergence of a potentially more transmissible variant of SARS-CoV-2, there is a need to better understand the dynamics of within-host accumulation of mutations in immunocompromised hosts, whether or not they remain infectious, if these mutations could mediate immune escape, and what kind of immunological selection pressures may drive such mutations. Altogether, these findings have important public health implications for infection control precautions and management in this patient population.

