## Supplementary Appendix for "Persistent SARS-CoV-2 infection and increasing viral variants in children and young adults with impaired humoral immunity"

**_­_Supplementary methods**

**ELISA to detect anti-SARS-CoV-2 antibodies in serum samples**

IgA, IgG and IgM to SARS-CoV-2 nucleocapsid (N) protein, spike S1, and receptor binding domain (RBD) and the were measured with a laboratory-developed ELISA as previously described by Röltgen et al.^1^ Briefly, 96-well Corning Costar high binding plates (Thermo Fisher) were coated with viral antigens at 0.1 μg per well (0.025 μg per well for the nucleocapsid IgG assay) overnight at 4°C. Serum or plasma samples were incubated in coated plates at a 1:100 dilution. Specific antibodies were detected with horseradish peroxidase conjugated goat anti-human IgG (γ-chain specific, catalog no. 62-8420, Thermo Fisher, 1:5,000 dilution), IgM (μ-chain specific, catalog no. A6907, Sigma, 1:5,000 dilution), or IgA (α-chain specific, catalog no. P0216, Agilent, 1:2,000 dilution). Plates were developed with 3,3′,5,5′-Tetramethylbenzidine (TMB) substrate and optical density (OD) at 450 nm was measured for sample wells with subtraction of blank well values. Seroconversion was defined as values above the mean plus three times the standard deviation of ELISA ODs from 94 pre-pandemic negative control samples from healthy blood donors. Control data were from non-immunocompromised COVID-19 patients less than 60 years old, sampled at least 50 days after symptom onset, and admitted to hospital without ICU care. All samples were tested twice in independent experiments.

**Competition ELISA to detect antibodies that block binding of ACE2 to RBD**

The competition ELISA was recently described in Röltgen et al.^1^ Briefly, RBD-coated ELISA plates were incubated with plasma samples at a 1:10 dilution before addition of recombinant ACE2 expressed as a fusion protein with mouse IgG2a Fc (ACE2-mFc) at 0.5 µg/mL. Detection of RBD-ACE2-mFc was done with horseradish peroxidase conjugated goat anti-mouse IgG (Fc specific, catalog no. 31439, Invitrogen, 1:10,000 dilution), developed and measured as described aboce. Two quality controls (Access SARS-CoV-2 IgG QC, QC1-QC2, 2 levels, catalog no. C58964, Beckman Coulter) were included on each plate. OD values were converted to percentage of RBD-ACE2 blocking using the following formula: percentage blocking = 100*(1-(sample OD - 0.2)/(QC1 OD – 0.2)), taking into account the background noise of the assay of 0.2 which was determined by testing historical negative control serum samples. All samples were tested three times in independent experiments.

**AmpliSeq multiplex-PCR method and S5XL sequencing:**

Confirmatory SARS-CoV-2 sequencing using the AmpliSeq SARS-CoV-2 Research Panel was done for viral samples with lower viral loads (Table S4). Viral RNA amplification and library preparation was performed according to the manufacturer’s protocol using the Ion AmpliSeq Library Kit Plus (Thermo Fisher Scientific) and two pools of custom primers. cDNA was synthesized using SuperScript VILO cDNA Synthesis Kit (SuperScript IV), in either 15 µL or 6 µL reactions using 9.6 or 4.8µL of RNA combined with 2.4 or 1.2 µL (respectively) of 5X VILO IV Master Mix (an additional 3µL of nuclease-free water was added to the 15 µL reactions); larger cDNA reactions were used for automated library prep on Ion Chef (Thermo Fisher Scientific) and the smaller volume reaction for manual library preparation. The reaction was incubated at 25°C for 10 minutes, 50°C for 10 minutes, and 85°C for 5 minutes. Following cDNA conversion, for manual library prep, separate multiplexed PCR reactions were set up for primer pool 1 and 2 using half of the cDNA for each primer pool, 2 µL of 5X Ion AmpliSeq HiFi Mix, 2 µL of 5X Ion AmpliSeq SARS-CoV-2 Research Panel primer pool 1 or 2, and nuclease-free water for a total volume of 10 µL per primer pool. PCR conditions used were as follows: initial activation of enzyme at 99°C for 2 minutes, 16-26 cycles of denaturation (99°C for 15 seconds) and annealing/extension (60°C for 4 minutes), and hold at 10°C. Primer pool 1 and 2 target amplification reactions were combined and partial digestion of amplicons and barcode ligation carried out according to the manufacturer’s protocol. The barcoded libraries then were purified using 1X beads-to-sample ratio of Agencourt AMPure XP Magnetic Beads and 70% Ethanol. The libraries were quantified using the Ion Library TaqMan Quantitation Kit and diluted to 30 pmol/L and pooled together. For Ion Chef automated library preparation, primer pools were diluted to 2x and added to the reagent cartridge; reagents, consumables, and cDNA in an IonCode PCR plate were loaded into the Ion Chef instrument per the Ion AmpliSeq on Chef protocol. A total of 16-27 cycles of amplification were performed for libraries generated on Chef with an anneal/extension time of 4 minutes. Final library pools generated by the Chef were quantified using the Ion Library TaqMan Quantitation Kit and normalized to 30 pmol/L.

A total of 25 µL of the pooled libraries was loaded onto the Ion Chef System for automated clonal amplification by emulsion PCR using Ion 510, 520/530 Chef Reagents and consumables and subsequent chip loading (520 or 530 chips). Viral genome sequencing was performed on the Ion Torrent S5 XL platform using standard 200-base sequencing chemistry.

**TWIST target-capture method and Illumina sequencing**

Confirmatory SARS-CoV-2 sequencing using the Twist SARS-CoV-2 Research Panel was done for viral samples with higher viral loads (Table S4). cDNA libraries were generated using Twist Library Preparation Kit for ssRNA Virus Detection. cDNA was generated per the Twist SARSCoV2-Virus Detection Protocol by annealing 5 µL Random Primer 6 (New England Biolabs) to up to 15 µL of RNA and incubating the 20 µL reaction volume at 95°C for 5 minutes; 25 µL of ProtoScript II Reaction Mix and 5 µL of ProtoScript II Enzyme Mix (New England Biolabs) were added to the primer annealed RNA for first strand synthesis (25°C for 5 minutes, 42°C for 60 minutes, and 80°C for 5 minutes). The NEBNext Ultra II Non-Directional RNA Second Strand Synthesis Module (New England Biolabs) was used to create second strand cDNA per the Twist protocol, and the cDNA was purified using SPRI Beads (Beckman Coulter Life Sciences). cDNA (enzymatic) fragmentation, end repair, and dA-tailing were carried out using the Twist Library Preparation EF Kit (Twist Bioscience) to generate TruSeq-compatible libraries. Universal adapters (Twist Biosciences) were ligated to libraries, and libraries were purified and subsequently amplified using Twist UDI primers and Kapa HiFi HotStart Ready Mix (Roche). A total of 12 cycles of amplification were used (98°C for 45 seconds, followed by 12 cycles of 98°C for 15 seconds, 60°C for 30 seconds, 72°C for 30 seconds, and a final extension of 72°C for 1 minute, and final hold at 4°C). Finally, the amplified, indexed libraries were purified and quantified using the Promega QuantiFluor ONE dsDNA System. A total of 1,500ng of each amplified product was hybridized to the Twist SARS-CoV-2 probe mix for 15 minutes at 60°C following the Twist protocol. Capture was performed using Twist binding beads and the Twist Fast Hybridization and Wash kit per the kit protocol. Captured libraries were amplified for 15 cycles (98°C for 45 seconds, followed by 15 cycles of 98°C for 15 seconds, 60°C for 30 seconds, 72°C for 30 seconds, and a final extension of 72°C for 1 minute, and final hold at 4°C). Amplified libraries were purified before library fragment profiles were quantified using the Agilent 4200 TapeStation High Sensitivity DNA assay. Library concentrations were normalized to ~5nM and pooled to a final pool concentration of 4nM then denatured and diluted according to the Illumina MiSeq Denature and Dilute Libraries Guide and loaded on the sequencer at 10pM. Paired-end and dual-indexed 2x150bp sequencing was done using Micro Kit v2 (300 Cycles).

**Consensus genome comparison, phylogenetic analysis and clade analysis**

Consensus genomes obtained on the patient isolates were compared to the Wuhan isolate (NC_045512.2) using SARS-CoV-2 Genome App v1.1 (https://cov2annot.cpmbiodev.net) and CHLA COVID-19 Analysis Research Database (CARD) to identify synonymous, non-synonymous and intergenic variations.^2,3^ Phylogenetic analysis and evolutionary rate estimation were performed using packages available through Nextstrain command-line interface (v 2.0.0.).^4^ Consensus viral sequences for each patient were combined to generate a multiple sequence alignment with MAFFT (version 7.460) using speed-oriented option - FFT-NS-i (iterative refinement method, two cycles) optimized for large datasets.^5^ A maximum likelihood tree using Bayesian information criteria was generated with IQ-TREE (version 2.0.3) using GTR substitution model.^6^ The resulting rate estimation and phylogeny was then time-resolved using TreeTime (version 0.7.6) and visualized using auspice.^4,7^ Phylogenetic clade analysis was performed using Nextclade (version 0.3.7).

**Supplementary figure**

**Figure S1.** A time-resolved evolutionary rate estimation for SARS-CoV-2 in patients with prolonged viral infectivity (patient 2 and patient 3). Collection date for each sample is shown on the x-axis. For patient 2, the estimated evolutionary rate was 2.01 x 10^-4^ substitutions per site. For patient 3, the estimated evolutionary rate was 1.38 X 10^-3^. The mutation rate is calculated based on differences in major and not minor variants, relative to the consensus difference.

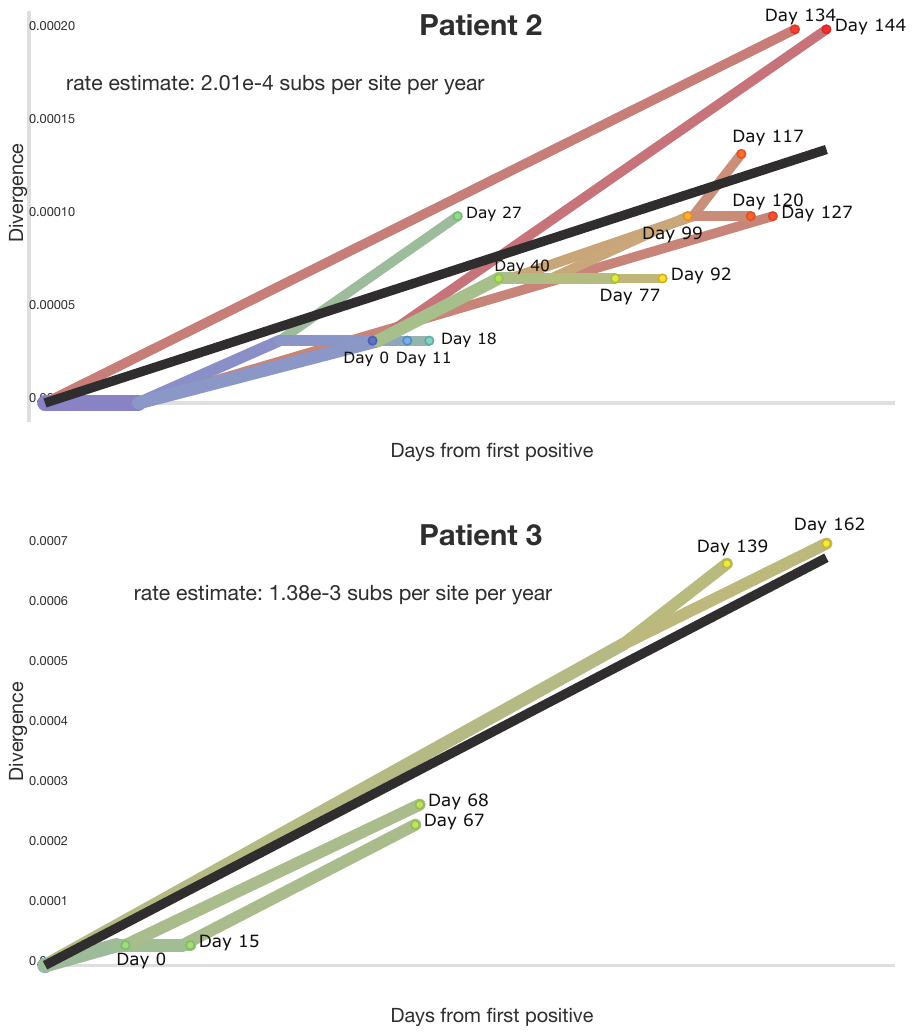

**Supplementary tables**

**Table S1.** RT-PCR assay values for detection of SARS-CoV-2 from nasopharyngeal or combined nares/oropharyngeal swabs collected from each patient. Corresponding viral culture assay results are also shown.

| Patient | Day | Routine RT-PCR | | Strand-specific RT-PCR | | Subgenomic RT-PCR  Ct | Viral culture |
| --- | --- | --- | --- | --- | --- | --- | --- |
|  |  | Assay used | Cycle threshold (Ct)^a^ | Minus strand Ct | Plus strand Ct |  |  |
| 1 | 0 | Thermo Fisher | 15.9 | 25.2 | 17.5 | 22.9 | Positive |
|  | 13 | Thermo Fisher | 26.2 |  |  |  | Negative |
|  | 27 | CDC | 33.3 |  |  |  |  |
|  | 34 | Thermo Fisher | 29.0 | UND | 34.1 | UND |  |
|  | 40 | Thermo Fisher | 30.7 | UND | 34.1 | UND |  |
|  | 46 | Thermo Fisher | 30.7 | UND | 35.1 | UND | Negative |
|  | 91 | Cepheid | UND |  |  |  |  |
|  | 116 | Thermo Fisher | UND | UND | UND | UND | Negative |
|  | 117 | Thermo Fisher | 37.5 | UND | UND | UND | Negative |
|  | 145 | Thermo Fisher | UND |  |  |  |  |
| 2 | 0 | CDC | 20.7 | 31.7 | 20.2 | 26.2 | Positive |
|  | 11 | Thermo Fisher | 21.4 | 29.5 | 22.3 | 29.6 | Positive |
|  | 18 | Thermo Fisher | 16.6 | 23.9 | 17.6 | 23.6 |  |
|  | 27 | Thermo Fisher | 16.2 |  |  |  | Positive |
|  | 40 | Thermo Fisher | 26.1 |  |  |  |  |
|  | 53 | Diasorin | 23.4 / 23.4 |  |  |  |  |
|  | 65 | Diasorin | 23.1 / 22.8 | 35.3 | 24.6 | 33.8 |  |
|  | 77 | Thermo Fisher | 21.8 |  |  |  |  |
|  | 92 | Thermo Fisher | 17.7 | 28.5 | 19.0 | 25.7 |  |
|  | 99 | Thermo Fisher | 20.2 | 29.5 | 21.8 | 27.8 | Positive |
|  | 117 | Thermo Fisher | 19.3 |  |  |  | Positive |
|  | 120 | Thermo Fisher | 18.5 | 27.5 | 20.8 | 25.6 | Positive |
|  | 127 | Thermo Fisher | 28.1 | UND | 27.7 | 36.8 | Positive |
|  | 130 | Thermo Fisher | 23.6 |  |  |  |  |
|  | 134 | Thermo Fisher | 20.9 | 28.9 | 22.5 | 29.3 | Positive |
|  | 137 | Thermo Fisher | 17.2 | 24.1 | 19.8 | 23.4 | Positive |
|  | 144 | Thermo Fisher | 21.6 | 30.7 | 24.3 | 31.2 | Positive |
|  | 156 | Thermo Fisher | 24.8 |  |  |  |  |
|  | 172 | Thermo Fisher | 23.1 |  |  |  |  |
| 3 | 0 | CDC | 16.0 | 23.4 | 17.5 | 25.0 | Positive |
|  | 11 | CDC | 22.5 |  |  |  |  |
|  | 15 | Thermo Fisher | 23.5 | 39.3 | 25.5 | 31.9 | Negative |
|  | 22 | Thermo Fisher | 21.8 | 32.1 | 24.2 | 32.9 | Positive |
|  | 36 | Diasorin | 19.8 / 19.2 |  |  |  |  |
|  | 43 | Diasorin | 21.2 / 22.3 |  |  |  |  |
|  | 67 | Thermo Fisher | 19.2 | 26.7 | 20.8 | 27.0 | Negative |
|  | 68 | Thermo Fisher | 21.9 | 30.7 | 22.8 | 31.4 |  |
|  | 139 | Thermo Fisher | 23.0 | 36.7 | 33.5 | 33.0 | Positive |
|  | 162^b^ | Thermo Fisher | 20.0 | 35.8 | 23.0 | 27.4 | Negative |
|  | 196 | Thermo Fisher | UND |  |  |  |  |

UND, undetected

^a^ Ct values for the Thermo Fisher Taqpath COVID-19 RT-PCR N target, CDC 2019-Novel Coronavirus Real-Time RT-PCR N1 target, Cepheid Xpert Xpress SARS-CoV-2 E and N2 targets, and Diasorin Simplexa S and ORF1ab targets.

^b^ All specimens were collected with a nasopharyngeal swab except for the day 162 specimen from patient 3, which was collected with a combined nares and oropharyngeal swab.

**Table S2. Major and minor allele frequency variants in patient 1.** Variants with allele frequency below 25% were filtered out. Most samples were processed on an additional platform to verify the variant calls. Variant calls confirmed by another platform are marked in bold. Patient 1, Day 27 sample with a low depth of coverage from Paragon run was replaced with AmpliSeq data. PAF = population allele frequency of variant in over 80,000 complete genome sequences in the Children’s Hospital Los Angeles COVID-19 Analysis Research Database (CARD), extracted from and kept updated based on GIDAID, NCBI, and Nextstrain.^3^

| Variant | | Gene | Day | | | PAF |
| --- | --- | --- | --- | --- | --- | --- |
|  |  |  | 0 | 13 | 27 |  |
| 241:C/T | . | 5UTR_orf1ab | **1** | **1** | **0.99** | 0.001 |
| 1059:C/T | p.Thr265Ile | orf1ab | **0.99** | **1** | **0.99** | 0.48 |
| 3037:C/T | p.Phe924Phe | orf1ab | **1** | **1** | **0.96** | 0.916 |
| 3042:C/T | p.Pro926Leu | orf1ab | **1** | **0.99** | **1** | <0.001 |
| 9967:C/T | p.Leu3234Leu | orf1ab | **1** | **1** | **1** | 0.002 |
| 11514:C/T | p.Thr3750Ile | orf1ab | **1** | **.** | **.** | 0.001 |
| 14408:C/T | p.Pro4715Leu | orf1ab | **1** | **1** | **0.99** | 0.916 |
| 21777:G/A | p.Gly72Glu | S | **1** | **1** | **0.98** | <0.001 |
| 22792:C/A | p.Ile410Ile | S | **1** | **1** | **0.49** | <0.001 |
| 23403:A/G | p.Asp614Gly | S | **1** | **1** | **1** | 0.920 |
| 24770:G/T | p.Ala1070Ser | S | **1** | **1** | **0.99** | <0.001 |
| 25563:G/T | p.Gln57His | ORF3a | **1** | **0.99** | **0.99** | 0.221 |
| 27964:C/T | p.Ser24Leu | ORF8 | **1** | **1** | **1** | 0.042 |
| 28824:C/T | p.Ser184Phe | N | **1** | **1** | **0.99** | <0.001 |

**Table S3. Major and minor allele frequency variants in patient 2.** Variants with allele frequency below 25% were filtered out. Most samples were processed on an additional platform to verify the variant calls. Variant calls confirmed by another platform are marked in bold. PAF = population allele frequency of variant in over 80,000 complete genome sequences in the Children’s Hospital Los Angeles COVID-19 Analysis Research Database (CARD), extracted from and kept updated based on GIDAID, NCBI, and Nextstrain.^3^

| **Variant** | | **Gene** | **Day** | | | | | | | | | | | | | | **PAF** |
| --- | --- | --- | --- | --- | --- | --- | --- | --- | --- | --- | --- | --- | --- | --- | --- | --- | --- |
|  |  |  | 0 | 11 | 18 | 27 | 40 | 77 | 92 | 99 | 117 | 120 | 127 | 134 | 137 | 144 |  |
| 241:C/T | . | 5UTR_orf1ab | **0.99** | **1** | **1** | **1** | **1** | **1** | **1** | **1** | **1** | **1** | **1** | **1** | 1 | 1 | 0.889 |
| 498:C/T | p.Thr78Ile | orf1ab | . | . | . | . | . | . | . | . | . | . | . | . | . | 0.47 | <0.001 |
| 509:G/A | p.Gly82Ser | orf1ab | . | . | . | . | . | . | . | . | **0.74** | **0.39** | . | . | . | . | <0.001 |
| 521:G/A | p.Val86Ile | orf1ab | . | . | . | . | . | . | . | **0.47** | . | . | . | . | . | . | <0.001 |
| 2939:C/G | p.Pro892Ala | orf1ab | . | . | . | . | . | . | . | . | . | . | . | **0.95** | . | 0.48 | <0.001 |
| 3037:C/T | p.Phe924  Phe | orf1ab | **1** | **1** | **1** | **1** | **1** | **1** | **1** | **1** | **0.99** | **1** | **0.99** | **1** | 1 | 1 | 0.916 |
| 3523:A/G | p.Glu1086  Glu | orf1ab | . | . | . | . | . | . | . | . | . | . | **0.43** | . | 0.75 | . | <0.001 |
| 7315:T/C | p.Phe2350  Phe | orf1ab | **1** | **1** | **1** | **1** | **1** | **1** | **1** | **1** | **1** | **1** | **1** | **1** | 1 | 0.99 | <0.001 |
| 8290:C/T | p.Leu2675  Leu | orf1ab | . | . | . | . | . | . | . | . | . | . | . | . | . | 0.57 | 0.001 |
| 8327:C/T | p.Leu2688  Phe | orf1ab | . | . | . | . | . | . | . | . | **0.33** | . | . | . | . | . | 0.001 |
| 9711:C/T | p.Ser3149  Phe | orf1ab | . | . | . | . | . | . | . | . | . | . | . | **0.95** | . | 0.44 | <0.001 |
| 10029:C/T | p.Thr3255  Ile | orf1ab | . | . | . | . | . | . | . | . | . | . | **0.64** | . | 0.99 | . | 0.001 |
| 10319:C/T | p.Leu3352  Phe | orf1ab | . | . | . | . | . | . | . | **0.53** | . | . | . | . | . | . | 0.024 |
| 10834:C/T | p.Ala3523  Ala | orf1ab | . | . | . | **0.61** | . | . | . | . | . | . | . | . | . | . | 0.001 |
| 11230:G/T | p.Met3655Ile | orf1ab | . | . | . | . | **0.99** | **0.78** | **0.99** | **0.96** | **0.93** | **0.99** | **0.36** | . | . | 0.58 | 0.009 |
| 12008:C/T | p.Leu3915  Phe | orf1ab | . | . | . | . | . | . | **0.40** | **0.28** | **0.85** | **0.94** | 0.25 | . | . | . | <0.001 |
| 14313:T/C | p.Asp4683  Asp | orf1ab | . | . | . | . | . | . | . | . | . | . | . | **0.72** | . | 0.43 | 0.001 |
| 14408:C/T | p.Pro4715  Leu | orf1ab | **0.99** | **1** | **1** | **1** | **1** | **1** | **1** | **1** | **1** | **1** | **1** | **1** | 1 | 0.99 | 0.916 |
| 14809:C/T | p.Arg4849  Cys | orf1ab | . | . | . | . | **0.98** | **0.74** | **0.98** | **0.96** | **0.92** | **0.99** | **0.36** | . | . | . | <0.001 |
| 15828:A/C | p.Glu5188  Asp | orf1ab | . | . | . | . | . | . | . | . | **0.37** | **0.28** | . | . | . | . | <0.001 |
| 16877:C/T | p.Thr5538Ile | orf1ab | . | . | . | . | . | . | . | . | . | . | . | . | . | 0.59 | <0.001 |
| 20384:C/T | p.Ala6707  Val | orf1ab | . | . | . | **0.46** | . | . | . | . | . | . | **0.68** | . | 0.98 | . | <0.001 |
| 21627:C/T | p.Thr22Ile | S | . | . | . | . | . | . | . | . | . | . | . | . | . | 0.60 | 0.001 |
| 21846:C/T | p.Thr95Ile | S | . | . | . | . | . | . | . | . | . | . | **0.51** | . | 0.76 | . | 0.002 |
| 21981:TTTTGGGTGTTTA/T^a^ | p.Leu141_Tyr144del | S | . | . | . | . | . | . | . | . | . | . | . | . | 0.78 | . | <0.001 |
| 21990:TTTA/T^a^ | p.Tyr145del | S | . | . | . | . | . | . | . | . | . | **0.43** | **0.33** | **0.97** | . | 0.44 | <0.001 |
| 22193:AATT/A | p.Asn211_  Leu212delinsIle | S | . | . | . | . | . | . | . | . | . | . | **0.57** | . | 0.95 | . | <0.001 |
| 22264:C/T | p.Asn234Asn | S | . | . | . | . | . | . | . | . | . | . | . | . | . | 0.58 | 0.001 |
| 23403:A/G | p.Asp614Gly | S | **1** | **1** | **1** | **1** | **1** | **1** | **1** | **1** | **1** | **1** | **1** | **1** | 1 | 1 | 0.920 |
| 24138:C/A | p.Thr859Asn | S | . | . | . | . | . | . | . | . | **0.34** | **0.29** | . | . | . | . | 0.001 |
| 25904:C/T | p.Ser171Leu | ORF3a | . | . | . | . | . | . | . | . | . | . | . | **0.76** | . | 0.36 | 0.004 |
| 26511:T/C |  | E-M | . | . | . | . | . | . | . | . | . | . | **0.49** | . | 0.98 | . | <0.001 |
| 27112:G/C | p.Ser197Thr | M | . | . | . | . | . | . | . | . | . | . | . | **0.53** | . | 0.29 | <0.001 |
| 27707:C/T | p.Ala105Val | ORF7a | . | . | . | . | **0.94** | **0.99** | **1** | **1** | **1** | **1** | **0.99** | **0.98** | 1 | 0.98 | 0.001 |
| 28321:G/A | p.Thr16Thr | N | . | . | . | . | . | **0.36** | . | . | . | . | . | **0.77** | . | 0.96 | 0.001 |
| 29679:C/T | . | ORF10_3UTR | . | . | . | . | . | . | . | . | **0.29** | . | . | . | . | . | <0.001 |
| 29700:A/T | . | ORF10_3UTR | . | . | . | . | . | . | . | . | . | . | . | . | 0.85 | . | <0.001 |
| 29742:G/T | . | ORF10_3UTR | . | . | . | **0.53** | . | . | . | . | . | . | . | . | . | . | 0.003 |

^a^ Leu141_Tyr144del and Tyr145del are in a region where similar mutations have been found in other immunocompromised patients.^8,9^

**Table S4. Major and minor allele frequency variants in patient 3.** Variants with allele frequency below 25% were filtered out. Most samples were processed on an additional platform to verify the variant calls. Variant calls confirmed by another platform are marked in bold. PAF = population allele frequency of variant in over 80,000 complete genome sequences in the Children’s Hospital Los Angeles COVID-19 Analysis Research Database (CARD), extracted from and kept updated based on GIDAID, NCBI, and Nextstrain.^3^

| Variant | | Gene | Day | | | | | | PAF |
| --- | --- | --- | --- | --- | --- | --- | --- | --- | --- |
|  |  |  | 0 | 15 | 67 | 68 | 139 | 162 |  |
| 241:C/T | . | 5UTR_  orf1ab | **1** | **1** | **1** | **1** | **1** | **1** | 0.889 |
| 379:C/A | p.Val38Val | orf1ab | **1** | **0.99** | **1** | **0.99** | **1** | **1** | 0.003 |
| 635:C/T | p.Arg124Cys | orf1ab | . | . | . | . | . | **0.44** | <0.001 |
| 1059:C/T | p.Thr265Ile | orf1ab | **1** | **0.99** | **0.99** | **0.99** | **1** | **1** | 0.148 |
| 1753:T/C | p.Tyr496Tyr | orf1ab | . | . | . | . | . | **0.47** | <0.001 |
| 2424:G/T | p.Cys720  Phe | orf1ab | . | . | . | . | . | **0.51** | <0.001 |
| 2655:A/C | p.Glu797  Ala | orf1ab | . | . | . | . | **1** | **1** | <0.001 |
| 3037:C/T | p.Phe924  Phe | orf1ab | **1** | **1** | **1** | **1** | **1** | **1** | 0.916 |
| 3421:A/G | p.Val1052  Val | orf1ab | . | . | . | . | **1** | **1** | <0.001 |
| 4763:C/T | p.His1500  Tyr | orf1ab | . | . | . | . | **1** | **1** | <0.001 |
| 4891:C/T | p.Thr1542  Thr | orf1ab | . | . | . | **0.62** | . | . | <0.001 |
| 9165:C/T | p.Thr2967Ile | orf1ab | . | . | . | . | **1** | **1** | 0.001 |
| 10276:A/G | p.Gln3337  Gln | orf1ab | . | . | . | . | . | **0.49** | <0.001 |
| 11082:TG/T | p.Leu3606fs | orf1ab | . | . | . | . | . | 0.27 | <0.001 |
| 11083:G/T | p.Leu3606  Phe | orf1ab | . | . | . | . | **0.47** | **0.73** | 0.065 |
| 11270:A/G | p.Met3669  Val | orf1ab | . | . | . | . | **1** | **1** | <0.001 |
| 12025:C/T | p.Ser3920  Ser | orf1ab | **1** | **1** | **1** | **1** | **1** | **1** | 0.002 |
| 13860:C/T | p.Asp4532  Asp | orf1ab | . | . | . | **0.62** | . | . | 0.001 |
| 14408:C/T | p.Pro4715  Leu | orf1ab | **0.99** | **1** | **1** | **1** | **1** | **1** | 0.916 |
| 15714:C/T | p.Leu5150  Leu | orf1ab | . | . | . | . | . | **0.51** | <0.001 |
| 16014:G/T | p.Arg5250  Arg | orf1ab | . | . | . | **0.59** | . | . | <0.001 |
| 16374:T/C | p.Asn5370  Asn | orf1ab | . | . | . | . | **0.99** | **0.51** | <0.001 |
| 16421:A/G | p.Gln5386  Arg | orf1ab | . | . | . | **0.41** | . | . | <0.001 |
| 16466:C/T | p.Pro5401  Leu | orf1ab | . | . | . | **0.44** | . | . | <0.001 |
| 16949:C/T | p.Pro5562  Leu | orf1ab | . | . | . | **0.60** | . | . | <0.001 |
| 17278:G/T | p.Val5672  Leu | orf1ab | . | . | . | **0.40** | . | . | 0.002 |
| 17895:T/C | p.Ala5877  Ala | orf1ab | . | . | . | . | **1** | **0.56** | <0.001 |
| 18318:G/T | p.Gly6018  Gly | orf1ab | . | . | . | . | . | **0.37** | <0.001 |
| 18870:T/A | p.Thr6202  Thr | orf1ab | . | . | . | . | **1** | **1** | <0.001 |
| 19398:G/T | p.Glu6378  Asp | orf1ab | . | . | **0.99** | . | . | . | <0.001 |
| 21600:G/T | p.Ser13Ile | S | . | . | . | . | **1** | **1** | 0.001 |
| 21658:C/T | p.Phe32Phe | S | . | . | . | **0.52** | . | . | 0.001 |
| 21770:G/T^a^ | p.Val70Phe | S | . | . | . | . | . | **0.42** | <0.001 |
| 21852:A/T | p.Lys97Met | S | . | . | . | . | **1** | **0.51** | <0.001 |
| 21980:TTTTT  GGGTG/T^c^ | p.Leu141_  Val143del | S | . | . | . | . | **0.99** | **0.99** | <0.001 |
| 22131:G/A | p.Arg190Lys | S | . | . | . | . | **1** | **1** | <0.001 |
| 22152:T/C | p.Ile197Thr | S | . | . | . | **0.58** | . | . | <0.001 |
| 22195:T/G | p.Asn211Lys | S | . | . | . | . | . | **0.56** | <0.001 |
| 22843:T/C | p.Asp427  Asp | S | . | . | . | **0.52** | . | . | <0.001 |
| 22882:T/A^b^ | p.Asn440Lys | S | . | . | . | . | . | **0.51** | <0.001 |
| 23010:T/C | p.Val483Ala | S | . | . | . | . | **1** | **1** | <0.001 |
| 23012:G/C | p.Glu484Gln | S | . | . | . | . | **1** | **0.45** | <0.001 |
| 23403:A/G | p.Asp614Gly | S | **1** | **1** | **1** | **1** | **1** | **1** | 0.920 |
| 23638:C/T | p.Ile692Ile | S | . | . | **0.97** | . | . | . | 0.001 |
| 24885:A/C | p.Asn1108  Thr | S | . | . | . | . | . | **0.39** | <0.001 |
| 25563:G/T | p.Gln57His | ORF3a | **1** | **0.99** | **0.99** | **1** | **1** | **1** | 0.221 |
| 26228:C/T | . | ORF3a-E | . | . | . | **0.43** | . | . | 0.001 |
| 26333:C/T | p.Thr30Ile | E | . | . | . | . | **1** | **0.59** | <0.001 |
| 26386:A/G | p.Asn48Asp | E | . | . | . | . | **1** | . | <0.001 |
| 26393:G/T | p.Ser50Ile | E | . | . | . | . | **0.96** | . | <0.001 |
| 26895:C/T | p.His125Tyr | M | . | . | . | . | **1** | **1** | 0.002 |
| 27393:C/T | . | ORF6-ORF7a | . | . | . | . | **1** | **1** | <0.001 |
| 27925:C/T | p.Thr11Ile | ORF8 | . | . | . | . | **1** | **0.89** | <0.001 |
| 28232:C/T | p.Asp113  Asp | ORF8 | . | . | . | **0.32** | . | . | <0.001 |
| 28253:C/T | p.Phe120  Phe | ORF8 | . | . | **0.99** | . | . | . | 0.005 |
| 28255:T/C | p.Ile121Thr | ORF8 | **0.83** | **0.84** | . | **0.77** | **0.86** | **0.79** | <0.001 |
| 28300:G/T | p.Gln9His | N | . | . | **0.99** | . | . | . | 0.003 |
| 28715:A/G | p.Thr148Ala | N | . | . | . | . | . | **0.59** | <0.001 |
| 28887:C/T | p.Thr205Ile | N | **0.96** | **0.74** | **1** | **0.61** | **1** | **1** | 0.008 |

^a^ Val70Phe overlaps the ∆69-70 deletion which has been reported in the B.1.1.7 variant and has been associated with other receptor binding domain changes.^10^

^b^ Asn440Lys was also reported by Weisblum et al. and Liu et al. by passaging SARS-CoV-2 in the presence of a monoclonal antibody. This mutation also emerged in an adult immunocompromised patient reported in Choi et al.^8,11,12^

^c^ Leu141_Val143del is in a region where similar mutations have been found in other immunocompromised patients.^8,9^

**Table S5. Confirmation of variant calls by alternate whole genome sequencing methods** Primary Paragon protocol was confirmed by either Ampliseq or Twist protocols.

| Sample | Platform |
| --- | --- |
| Patient 1, Day 0 | Twist |
| Patient 1, Day 13 | AmpliSeq |
| Patient 1, Day 27 | n/a |
| Patient 2, Day 0 | Twist |
| Patient 2, Day 11 | Twist |
| Patient 2, Day 18 | Twist |
| Patient 2, Day 27 | Twist |
| Patient 2, Day 40 | AmpliSeq |
| Patient 2, Day 77 | Twist |
| Patient 2, Day 92 | Twist |
| Patient 2, Day 99 | Twist |
| Patient 2, Day 117 | Twist |
| Patient 2, Day 120 | Twist |
| Patient 2, Day 127 | AmpliSeq |
| Patient 2, Day 134 | Twist |
| Patient 2, Day 137 | n/a |
| Patient 2, Day 144 | n/a |
| Patient 3, Day 0 | Twist |
| Patient 3, Day 15 | AmpliSeq |
| Patient 3, Day 67 | Twist |
| Patient 3, Day 68 | Twist |
| Patient 3, Day 139 | AmpliSeq |
| Patient 3, Day 162 | Twist |
